# Evaluation of a multi-species SARS-CoV-2 surrogate virus neutralization test

**DOI:** 10.1101/2021.05.07.21252267

**Authors:** Carmen W.E. Embregts, Babs Verstrepen, Jan A.M. Langermans, Kinga P. Böszörményi, Reina S. Sikkema, Rory D. de Vries, Donata Hoffmann, Kerstin Wernike, Lidwien A.M. Smit, Shan Zhao, Barry Rockx, Marion P.G. Koopmans, Bart L. Haagmans, Thijs Kuiken, Corine H. GeurtsvanKessel

## Abstract

Assays to measure SARS-CoV-2-specific neutralizing antibodies are important to monitor seroprevalence, to study asymptomatic infections and to reveal (intermediate) hosts. A recently developed assay, the surrogate virus-neutralization test (sVNT) is a quick and commercially available alternative to the “gold standard” virus neutralization assay using authentic virus, and does not require processing at BSL-3 level. The assay relies on the inhibition of binding of the receptor binding domain (RBD) on the spike (S) protein to human angiotensin-converting enzyme 2 (hACE2) by antibodies present in sera. As the sVNT does not require species- or isotype-specific conjugates, it can be similarly used for antibody detection in human and animal sera. In this study, we used 298 sera from PCR-confirmed COVID-19 patients and 151 sera from patients confirmed with other coronavirus or other (respiratory) infections, to evaluate the performance of the sVNT. To analyze the use of the assay in a One Health setting, we studied the presence of RBD-binding antibodies in 154 sera from nine animal species (cynomolgus and rhesus macaques, ferrets, rabbits, hamsters, cats, cattle, mink and dromedary camels). The sVNT showed a moderate to high sensitivity and a high specificity using sera from confirmed COVID-19 patients (91.3% and 100%, respectively) and animal sera (93.9% and 100%), however it lacked sensitivity to detect low titers. Significant correlations were found between the sVNT outcomes and PRNT_50_ and the Wantai total Ig and IgM ELISAs. While species-specific validation will be essential, our results show that the sVNT holds promise in detecting RBD-binding antibodies in multiple species.

## Introduction

The severe acute respiratory syndrome coronavirus 2 (SARS-CoV-2) likely originates from an animal reservoir as a result of a direct spill-over event or via an intermediate mammalian host, similar to the related zoonotic betacoronaviruses SARS-CoV and MERS-CoV [1,2]. Phylogenetic analysis revealed that SARS-CoV-2 is ancestrally linked to betacoronaviruses found in bats [3] and pangolins [4], however, the definitive virus origin and intermediate host(s) remain unidentified. Besides efficiently infecting humans, SARS-CoV-2 has been detected in a wide range of animals, including farmed mink across Europe and the USA [5–7], domestic animals including cats and dogs [8–10], and several zoo felids [11]. Alarmingly, infections in all these species could be traced back to SARS-CoV-2 infected humans, indicating a risk for reverse zoonotic events and possible SARS-CoV-2 animal reservoirs [7,12]. Furthermore, infection experiments show that many more animal species, including non-human primates [13,14], ferrets [15,16], rabbits [17], hamsters [18,19], and human angiotensin-converting enzyme 2 (hACE2) transgenic mice [20] are permissive to the virus, while other animals including pigs and chickens are not [21,22]. The large number of permissive species and the potential risks of additional (reverse) zoonotic events clearly indicate that a One Health approach is required to gain insights into the circulation of SARS-CoV-2 in humans and epidemiologically connected animal host populations, which is essential for the prevention or mitigation of further spread.

Assays to reliably detect SARS-CoV-2 specific antibodies across species are urgently needed, for example to investigate seroprevalence and asymptomatic infections, for vaccination studies in humans and animals, and for the identification of natural reservoirs and intermediate hosts. Tremendous efforts in the rapid development of serological tools yielded a broad range of assays to determine SARS-CoV-2 specific antibodies, including (high throughput) ELISAs and lateral flow assays targeting various SARS-CoV-2 epitopes [23–25]. Total serum antibodies are indicative for exposure, however, quantifying neutralizing antibodies is more informative. A commonly used gold standard for detecting SARS-CoV-2 neutralizing antibodies is the 50-percent plaque-reduction neutralization test (PRNT_50_). This test requires handling of wild-type viruses by BSL3-level trained personnel, is not suitable for high-throughput, and results are available after multiple days. Furthermore, minor differences in virus stocks and cell lines complicate the intra-laboratory standardization. While pseudotyped viruses allow for performing PRNT_50_ tests at a BSL-2 safety level [26,27], and recombinant nanoluciferase SARS-CoV-2 allows for a rapid assay protocol [28], these assays still rely on infectious viruses and cell cultures.

The first surrogate virus neutralization test (sVNT) was commercialized in 2020 [29]. This assay relies on specific binding of recombinant SARS-CoV-2 receptor binding domain (RBD) to recombinant ACE2 coated on 96-wells plates, and blocking of this binding by RBD-specific serum antibodies. The assay can be performed at any BSL-2 laboratory and yields results in only few hours. Furthermore, the assay allows for high sample-throughput, as samples are analyzed in one defined dilution and no serial dilution is required. Validation studies showed high specificity and sensitivity of the assay [29–31], however, a recent study demonstrated low sensitivity in sera with low neutralizing titers and only moderate linearity with the PRNT_50_ [32]. While conventional ELISAs often rely on species- and isotype-specific conjugates, the sVNT assay detects RBD-binding antibodies and can potentially be used for a wide range of species. In contrast to the large number of tests developed for human sera, serological SARS-CoV-2 assays for other species are limited to the PRNT_50_, few multi-species ELISAs [16,33,34], and mouse-, guinea pig-, rabbit-, and primate-specific competition ELISAs [35,36].

To date, only few studies assessed the use of the described sVNT in animals (mice, rabbits, ferrets, cats and hamsters) [29,30]. While these studies showed that the sVNT is capable of detecting RBD-specific antibodies in the mentioned animal species, only limited numbers of SARS-CoV-2 antibody positive sera were included. Here, we evaluate the performance of the sVNT on human sera using 298 serum samples from a COVID-19 patient cohort, 151 sera from patients diagnosed with related human coronaviruses and other (respiratory) viruses and pathogens. In addition, we investigate the use of the sVNT on 154 serum samples from nine different animal species.

## Materials and methods

### Human serum samples

All human sera used in this study were collected for routine patient diagnostics. Sensitivity analysis was performed using a panel of 298 sera of 165 PCR-confirmed COVID-19 patients. Disease severity ranged from mild (non-hospitalized) to severe (admitted to the ICU), and samples were taken at various days post disease onset (dpd), ranging from 0-74 days (Supplementary table 1). Specificity analysis was performed using a panel of 151 sera from individuals exposed to other human coronaviruses (HCoV-229E (n=19), HCoV-NL63 (n=18), HCoV-OC43 (n=36), or MERS-CoV (n=5)), other respiratory viruses (adenovirus (n=6), bocavirus (n=2), human metapneumovirus (HMPV, n=9), influenza virus A (n=10) and B (n=6), human orthopneumovirus/ respiratory syncytial virus (RSV) A (n=5) and B (n=4), rhinovirus (n=9), para-influenza virus 1 (n=4) and 3 (n=4), enterovirus (n=2)), or patients with recent cytomegalovirus (CMV, n=4), Epstein Barr virus (EBV, n=7) or *Mycoplasma pneumoniae* (*M. pneumoniae*, n=1) infection. All sera were stored at -20 °C and were heat-inactivated at 56 °C for 30 minutes prior to analysis.

### Animal serum samples

Animal sera were obtained after natural infections or during infection or vaccination experiments. In total 154 sera of nine different species (cynomolgus and rhesus macaques, ferrets, rabbits, hamsters, cats, cattle, mink and dromedary camels) were included in the validation study, of which 66 tested positive for SARS-CoV-2 antibodies by PRNT_50_ (Supplementary table 2). SARS-CoV-2 antibody status of the cat sera was determined by a pseudotype VSV neutralization (VN) assay [37]. Sera from SARS-CoV-2 negative animals were included to test the specificity of the assay. A set of possible cross-reactive sera from MERS-CoV infected macaques and dromedary camels was included as well.

### Ethics

The use of human specimens was approved by the Erasmus MC medical ethical committee (MEC approval: 2014–414), which allows the use of clinical data and left-over material from the specimen delivered to our laboratory for diagnostics, unless patients have declared they opted out of this scheme.

Animal sera were obtained as left-over material from various infection experiments or field studies (mink and cats). Specific approval was obtained for each set of sera and can be found in the referred articles. Additional non-human primate sera were obtained from various experiments that were approved by the Dutch Central Committee for Animal Experiments (license: AVD5020020209404). Mink and cat sera were obtained by a certified veterinarian during a SARS-CoV-2 outbreak at a mink farm in the Netherlands.

### PRNT_50_ / VSV pseudotype VN

The 50-percent plaque-reduction neutralization test (PRNT_50_) was used as the gold standard in this study and was performed as described before [38]. The PRNT_50_ titer was defined as the reciprocal value of the highest serum dilution resulting in 50% plaque reduction. Serum titers of ≥ 20 were defined as SARS-CoV-2 seropositive.

The pseudotype VN assay was performed as described recently [37] with some minor modification: serum samples were twofold diluted (starting at 1:8) and mixed 1:1 with SARS2-VSV. Mixtures were pre-incubated at 37°C for one hour and were afterwards used for inoculation of cells. Twenty-four hours post infection, the cells were lysed and relative luminescence units (RLU) of luciferase activity was determined. The sample neutralization titers were defined by the reciprocal of the highest dilution that resulted in >50% reduction of luciferase activity (IC50 titer).

### Wantai Ig/IgM

Detection of anti-RBD antibodies was performed using the Wantai SARS-CoV-2 total Ig or IgM ELISAs (Beijing Wantai Biological Pharmacy Enterprise), which are sandwich ELISAs coated with recombinant RBD. The ELISAs were performed according to the manufacturer’s guidelines. The readout (OD ratio) was calculated by dividing the OD (measured at 450 nm) of each sample with the OD of the calibrator that was supplied with the kit.

### Surrogate VNT

RBD-binding antibodies in human and animal sera were measured with the GenScript cPass SARS-CoV-2 Neutralization Antibody Detection Kit (Genscript, the Netherlands), following the manufacturer’s guidelines. Briefly, serum samples (1:10 diluted) were mixed with equal volumes of recombinant HRP-conjugated RBD and incubated for 30 minutes at 37 °C. One hundred µL was then transferred to 96-well plates coated with recombinant hACE2 receptor and incubated for 15 minutes at 37 °C. The mixture was removed, and after four automated washing steps, the development solution (tetramethylbenzidine substrate, TMB) was incubated for 15 minutes at room temperature, after which the stop solution was added. Absorbance was measured at 450 nm and the percentage of inhibition of each sample was calculated using the following formula: % inhibition = (1- (OD450 sample/ OD450 of negative control)) x 100. Controls were included in duplicate, samples were analyzed in singular. Inhibition >30% was regarded as a positive neutralization, as suggested by the latest validation paper [29].

### Statistical analysis

Spearman’s correlation coefficients were calculated on the sVNT inhibition percentages and the log2-transformed PNRT_50_ titers or the ODratio for the Wantai Ig or IgM in SPSS 27 (IBM). Correlation was considered significant with *p* values < 0.05.

## Results and discussion

The performance of the commercial sVNT was evaluated by determining the correlation between the PRNT_50_, a gold standard assay, and the sVNT. Although the initial commercial sVNT guidelines included a cut-off of 20%, a recent validation paper now recommends a positivity cut-off at 30% of inhibition [29]. We therefore evaluated the performance of the sVNT by both a 20% and 30% cut-off (Table 1). For the discussion of our results, we will focus on the evaluation with the 30% cut-off. Using the serum panel of PCR-confirmed COVID-19 patients we found an overall sensitivity of 91.3 and a corresponding specificity of 100%. We found a strong increase in sensitivity of the assay with increasing PRNT_50_ titers; sensitivity rose from respectively 50% and 74.1% in the low-titer groups of 20 and 40, to 91.4% and above for titers of 80 and higher (Table 1, Fig. 1A). 100% sensitivity was reached for sera with titers of 160 and above. In line with the expected rise in titer during the course of disease, we found that assay sensitivity increased from 88.2 to 91.0 and 96.6 when comparing the periods between 1-10, 11-21 and >21 dpd (Table 1). Overall, a significant (*p* < 0.001, with an Spearman’s *r* of 0.68) correlation was observed between the two serological tests. However, the variation within PRNT_50_ groups shows that the sVNT results should be interpreted with care, since high inhibition in the sVNT is not directly translatable to high PRNT_50_ titers (Fig. 1A). Performing a 30%-inhibition titration of each sample would allow a more accurate comparison of both assays. However, this would drastically decrease sample throughput and increase costs, which is unpreferable in diagnostic settings.

**Table 1:**
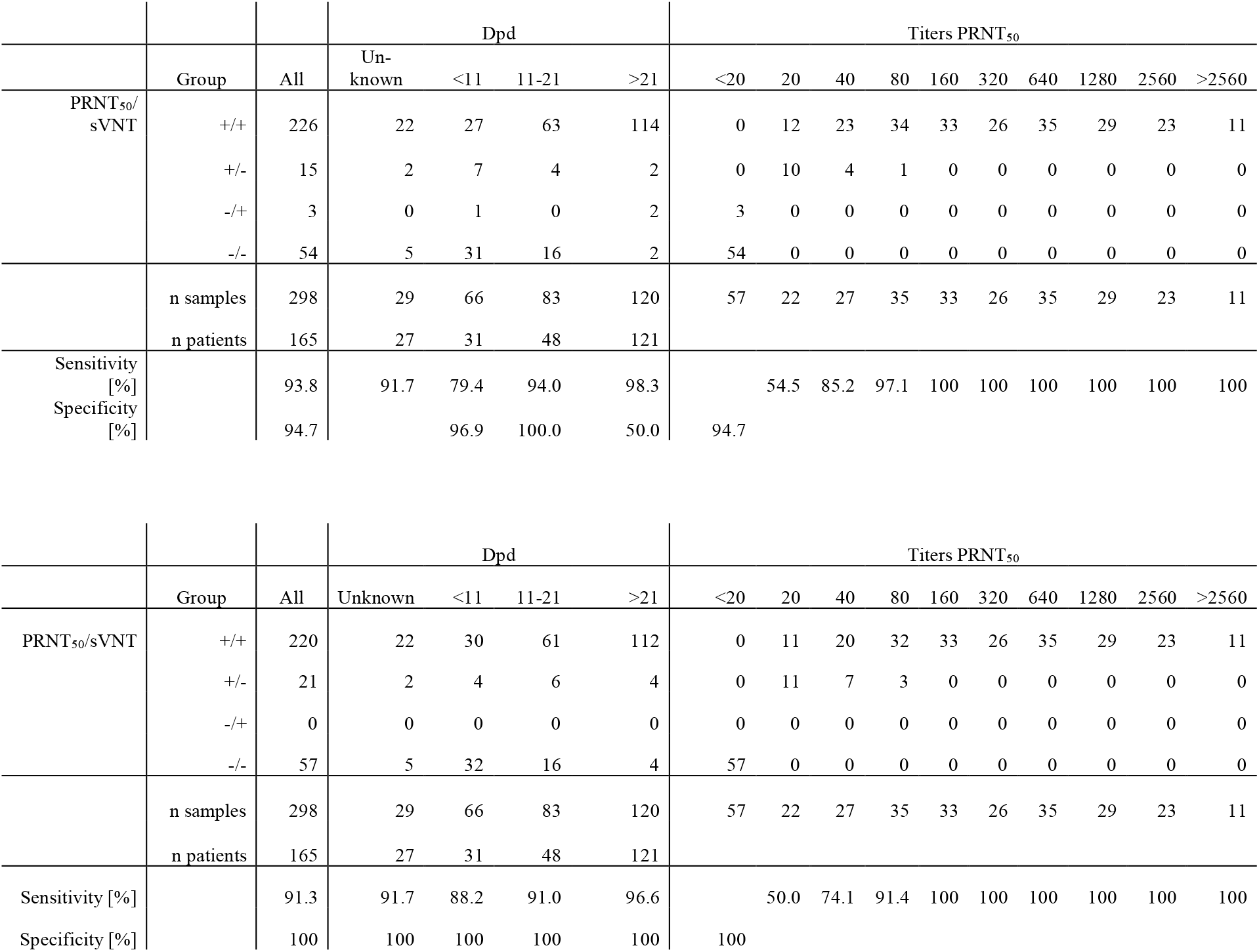
Sensitivity and specificity analysis of the sVNT using a 20% inhibition cut-off [upper part] or 30% cut-off of inhibition [lower part].

**Table 2.** Sensitivity and specificity analysis of the sVNT using an animal serum panel of nine different species, including SARS-CoV and MERS-CoV positive animals and control animals.

|  | SARS-CoV-2 |  |  |  |
| --- | --- | --- | --- | --- |
|  | Seropositive |  | Seronegative |  |
|  | 20% | 30% | 20% | 30% |
| Cynomolgus macaques | 11/13 | 9/13 | 10/10 | 10/10 |
| Rhesus macaques | 9/9 | 9/9 | 10/10 | 10/10 |
| Ferrets | 16/16 | 16/16 | 11/11 | 11/11 |
| Rabbits | 7/7 | 7/7 | 14/14 | 14/14 |
| Hamsters | 4/4 | 4/4 | 4/4 | 4/4 |
| Cats | 6/6 | 6/6 | 9/10 | 10/10 |
| Cattle | 1/1 | 1/1 | 10/10 | 10/10 |
| Mink | 10/10 | 10/10 | - | - |
| MERS-CoV-seropositive |  |  |  |  |
| Dromedary camels |  |  | 13/13 | 13/13 |
| Cynomolgus macaques |  |  | 2/2 | 2/2 |
|  | Sensitivity |  | Specificity |  |
|  | 20% | 30% | 20% | 30% |
| All data | 97.0 | 93.9 | 99.07 | 100 |
| PRNT<sub>50</sub> 20-80 | 92.9 | 85.7 |  |  |
| PRNT<sub>50</sub> >80 | 100 | 100 |  |  |

**Fig. 1.**
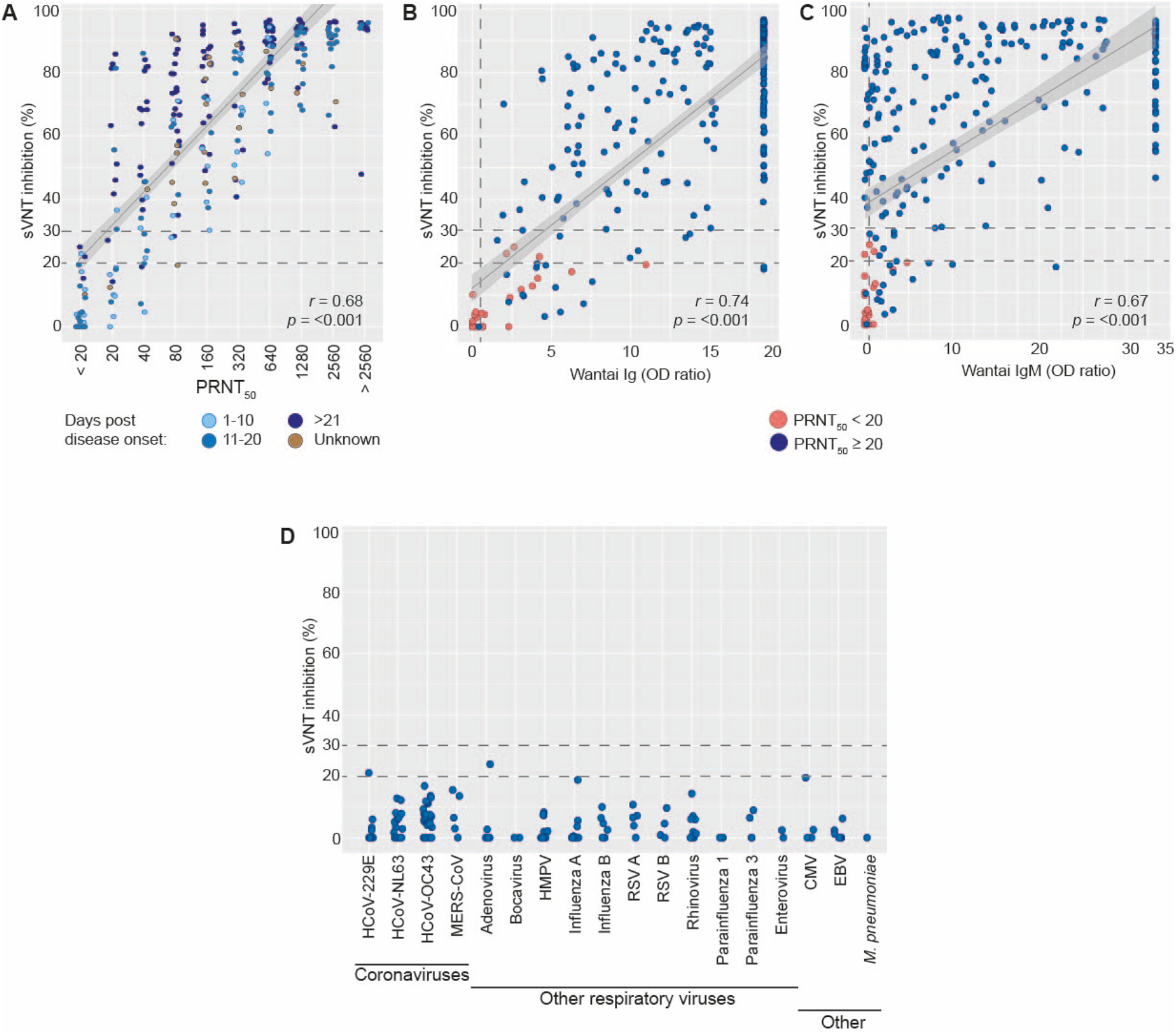
sVNT results using the human validation panel containing confirmed COVID-19 patients compared to the PRNT_50_ (A), the total Ig Wantai ELISA (B) or the IgM Wantai ELISA (C). sVNT results using the cross-reactive human serum panel are shown in (D). Each dot represents an individual serum. Dotted grey lines indicate the suggested cut-off values for the presented assays. The grey lines in (A), (B) and (C) show the Spearman’s correlation coefficient (*r*) and its confidence interval (grey area) of the PRNT_50_ / Wantai ELISAs and the sVNT.

The results of the sVNT also showed significant correlation (*p* < 0.001) with OD ratios of the Wantai SARS-CoV-2 specific total Ig or IgM ELISAs, with a Spearman’s *r* of 0.74 and 0.67 for the total Ig (Fig. 1B) and IgM (Fig. 1C) Wantai ELISA, respectively. Unfortunately, the high number of samples that reached the maximum value in both ELISAs might have affected the correlation coefficient. Closer investigation of the sera that showed a positive PRNT_50_ result but a negative sVNT result revealed that 18 out of 21 sera were positive in the IgM Wantai and 20 out of 21 were positive in the total Ig Wantai. SARS-CoV-2 neutralizing antibodies that do not block binding of RBD to ACE2 have been described [37], suggesting that this type of antibodies might cause false negative results in the sVNT. Another risk for false negatives is that this sVNT only targets RBD-binding antibodies, leaving neutralizing antibodies against other domains of the S1-protein undetected [39,40].

The specificity of the sVNT was further investigated using a serum panel containing sera of individuals diagnosed with other coronaviruses or other (respiratory) viruses or diseases. Using this panel, we confirmed that the sVNT is 100% specific, as we did not find cross-reactivity with any of the tested sera (Fig. 1B). Two samples were found to have an inhibition between 20-30%, one serum of a HCoV-229E patient and one of an adenovirus patient. Both samples tested negative in the PRNT_50_.

In parallel to the human sera, we assessed the performance of the sVNT in an elaborate panel of animal sera that included experimental model species, but also (suspected) reservoir species (Fig. 2A). Specificity of the assay was assessed using a panel of control sera from naïve animals. Sera from MERS-CoV infected cynomolgus macaques and dromedary camels were included to assess possible cross-reactivity. Similar to the results obtained with the human validation serum panel, the sVNT showed a good performance in general, with a sensitivity of 93.9% and a specificity 100%. For the rhesus macaques, ferrets, rabbits, hamsters, cats, cattle and mink we observed a 100% accurate detection of (the absence of) RBD-specific antibodies in the sera of SARS-CoV-2 positive and negative animals.

**Fig. 2.**
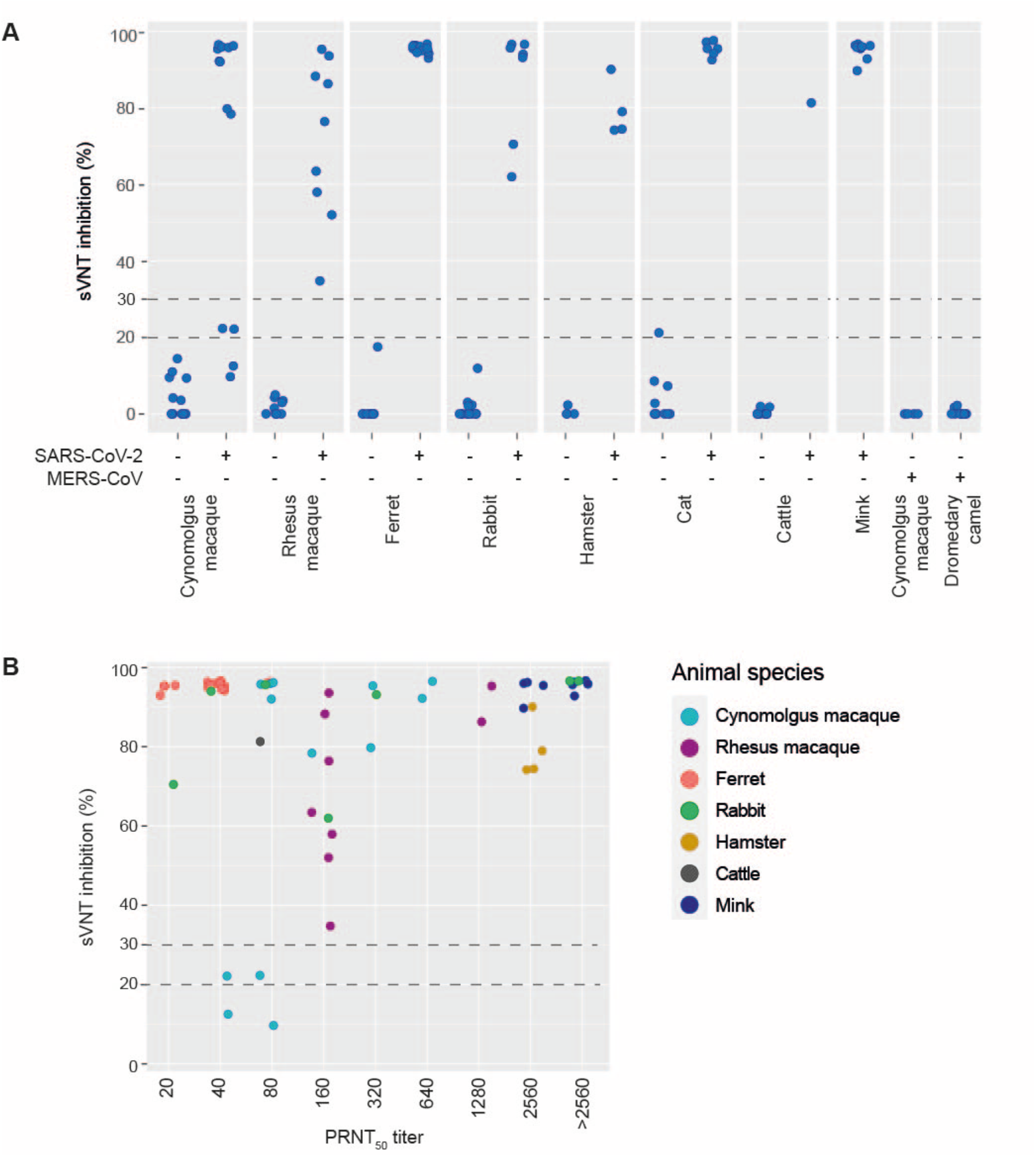
sVNT results obtained with sera of SARS-CoV-2-infected animals, MERS-CoV-infected animals or control animals, grouped by animal species and seroconversion status (positive vs. negative, (A)), or PRNT_50_ titer (B). Each dot represents an individual serum. Dotted horizontal lines indicate the suggested 20% and 30% inhibition cut-off values. Cats were excluded in (B) since no PRNT_50_ were available.

For the cynomolgus macaques two sera of SARS-CoV-2 infected animals were found to have an inhibition between 20% and 30%, and a PRNT_50_ titer of 40 and 80 (Fig. 2B). However, low antibody titers were expected since these animals only showed a short period of viral shedding with low levels of viral RNA in nose and trachea.

In agreement with the human cross-reactive serum panel, no cross-reactivity was detected in serum from MERS-CoV-infected rabbits and dromedary camels. The sVNT showed a sensitivity and specificity of 100% in animal sera with a PRNT_50_ titer of 160 and above. In contrast to the panel of human sera, no clear linearity was detected in the panel of animal sera and large differences were observed between species. Interestingly, serum samples from ferrets, rabbits and cattle with relatively low PRNT_50_ titers (80 and below) showed a high inhibition in the sVNT. Especially SARS-CoV-2-infected ferrets, where only two animals reached a PRNT_50_ titer of 80, had an inhibition of above 92%. While these high levels of inhibition in sera with a relatively low PRNT_50_ is beneficial for detecting RBD-binding antibodies in a qualitative manner, species-specific determination of the optimal serum dilutions is essential when the data is to be interpreted (semi-)quantitatively. The control sera of the ferrets, rabbits and cattle had negative sVNT outcomes, indicating that the high inhibition levels were not due to background or aspecific binding. Furthermore, the wide range of species-specific endemic coronaviruses complicates the design of specific serological tests [41,42], and cross-reactivity needs to be examined for every targeted species. While our data shows that the sVNT detects RBD-binding antibodies in nine animal species, it clearly indicates that more elaborate validation is required. Validations should include higher number of sera per species, a panel of potentially cross-reactive sera, and should aim at determining optimal serum dilutions and cut-off levels.

## Conclusion

Our results show moderate to high sensitivity and high specificity of the sVNT for detecting RBD-binding antibodies, with a 100% accuracy in sera with a PRNT_50_ titer 160, for both human and animal sera. sVNT results should be interpreted rather qualitatively than quantitatively, since the results only show partial linearity with the PRNT_50_ titers.

Despite the low sensitivity in detecting low titers, the sVNT still has potential use. The possibility for high sample throughput makes the sVNT a suitable assay for large seroprevalence studies that aim at detecting high titers, for example in vaccination trials or in large scale initial testing of potential animal reservoirs. While the required titer for complete protection is still under investigation, studies have shown that with a PRNT_50_ titer of 80 and above, no infectious virus could be detected in the respiratory tract [43]. It is thus to be expected that threshold titers for complete protection will be in this range or higher, and as a consequence the sVNT can be a valuable assay to assess protection in a qualitative manner. However, the sVNT does not serve as a full replacement of gold standard tests that use authentic virus, given that it lacks the sensitivity to detect low titers and only targets RBD-binding antibodies.

Our evaluation shows that the sVNT also has potential use for detecting RBD-specific antibodies in animal sera, but we observed large species-dependent differences in sensitivity of the test. While in some species we observed high sVNT results in sera with low PRNT_50_ (ferrets, rabbit, cattle), sera with low to moderate PRNT_50_ from other species resulted in negative or low sVNT results (cynomolgus macaques). More elaborate species-specific validations are required to determine the true potential of the sVNT.

## Data Availability

All data will be made available upon request.

## Acknowledgments

The authors acknowledge Anouskha Comvalius, Djenolan van Mourik, Jeroen Ijpelaar and Georgina Arron for performing the PRNT_50_, and Susanne Bogers for the sVNT testing. Robert Jan Molenaar is acknowledged for supplying the mink sera that was included in this study.

## Supplementary tables

**Supplementary table 1.** Overview of the human COVID-19 validation serum panel.

|  | N | Age | Sex |  |  | Dpd | Outcome |  |  |
| --- | --- | --- | --- | --- | --- | --- | --- | --- | --- |
|  |  | Mean [range] | Female | Male | Unknown | Mean [range]* | Discharged | Deceased | Unknown |
| All | 298 | 53 [15-90] | 61 | 216 | 21 | 26 [0-74] | 94 | 48 | 156 |
| Mild | 132 | 51 [15-66] | 25 | 88 | 19 | 25 [0-74] | 2 | 0 | 130 |
| Ward | 61 | 52 [25-90] | 14 | 45 | 2 | 25 [2-65] | 23 | 19 | 19 |
| ICU | 105 | 52 [17-83] | 22 | 83 | 0 | 24 [0-41] | 69 | 29 | 7 |

\* 29 samples had an unknown dpd

**Supplementary table 2.** Overview of sera included in the animal serum validation panel.

|  | SARS-CoV-2 | Control | Other | Total | Reference |
| --- | --- | --- | --- | --- | --- |
| Cynomolgus macaques | 13 | 14 | 2 [MERS-CoV] | 29 | [13, unpublished] |
| Rhesus macaques | 9 | 10 |  | 19 | unpublished |
| Ferrets | 16 | 11 |  | 27 | [36, unpublished] |
| Rabbits | 7 | 14 |  | 21 | [17,33] |
| Hamsters | 4 | 4 |  | 8 | [44] |
| Cats | 6 | 10 |  | 16 | [5] |
| Cattle | 1 | 10 |  | 11 | [45] |
| Mink | 10 | 0 |  | 10 | unpublished |
| Dromedary camels | 0 | 0 | 13 [MERS-CoV] | 13 | [46] |
|  | 66 | 73 | 15 | 154 |  |

